# A More Accurate Measurement of COVID-19 Hospitalizations

**DOI:** 10.1101/2022.06.03.22275891

**Authors:** Christina Vu, Eric S. Kawaguchi, Cesar H. Torres, Austin H. Lee, Chrysovalantis Stafylis, Jeffrey D. Klausner, Saahir Khan

**Affiliations:** Department of Medicine, Keck School of Medicine, University of Southern California, Los Angeles, California, USA; Department of Clinical Population and Public Health Sciences, Keck School of Medicine, University of Southern California, Los Angeles, California, USA; Keck School of Medicine, University of Southern California, Los Angeles, California, USA

**Author notes:** **Corresponding Author:** Saahir Khan, MD, PhD, Assistant Clinical Professor of Infectious Diseases, Keck School of Medicine of the University of Southern California, 2020 Zonal Ave., Rm. 433, Los Angeles, CA 90033.

**Keywords:** COVID-19, SARS-CoV-2, Hospital Epidemiology, Vaccination, Infection

## Abstract

COVID-19 hospitalizations are a key indicator of the SARS-CoV-2 pandemic. The US Centers for Disease Prevention and Control describes oxygen supplementation as a measure of severe disease. SARS-CoV-2 PCR positive patients hospitalized without hypoxia or requiring oxygen may have been identified incidentally due to routine screening practices. We describe the application of a revised case definition for COVID-19 hospitalization based on the case-patient oxygen requirement on admission. Using data collected from consecutive SARS-CoV-2 PCR positive hospital admissions in December 2021 and January 2022 at a large safety net hospital in Los Angeles County, we highlight differences between patients hospitalized *with* COVID-19 (i.e., no oxygen requirement on admission or `incidental` infection) and those hospitalized *for* COVID-19 (i.e., oxygen requirement on admission). We conducted multivariable modeling to determine the effect of age as a positive predictor of COVID-19 hospitalization and vaccination or prior infection as a negative predictor. The revised case-definition resulted in a substantial decrease in the number of COVID-19 hospitalizations during the study period: 67.5% of SARS-CoV-2 PCR positive hospital admissions were not *for* COVID-19 but *with* COVID-19. A revised case-definition for COVID-19 hospitalization that includes the oxygen requirement on admission is needed to more accurately monitor the pandemic and inform public health policy.

---

While preventing infection was the initial focus of the COVID-19 pandemic response, with increasing population immunity and variant transmissibility, the current focus has shifted to reducing hospitalization and deaths, particularly in vulnerable communities (1). During the recent surge in disease activity driven by the Omicron variant, an increased proportion of “COVID-19 hospitalizations” were incidentally discovered infections in patients newly hospitalized for other reasons (2-6), resulting in decreased measurements of in-hospital disease severity and mortality compared to prior disease surges (6-9). However, estimates of the proportion of total COVID-19 hospitalizations accounted for by these incidental infections range widely from 15% to 68% (2-6), due to heterogeneity in case definitions for these incidental infections and variability across populations with respect to vaccination status and other risk factors for severe COVID-19.

We propose utilizing the CDC criteria for severe COVID-19, based on need for supplemental oxygen or oxygen saturation measured below 92%, to define incidental COVID-19 hospitalization (10). To study the impact of this case definition, we reviewed medical records of SARS-CoV-2 PCR positive patients admitted to LAC+USC Medical Center, a safety net hospital serving predominantly Latino and low-income patients in Los Angeles, California, during the local Omicron variant surge between 12/10/21 to 1/19/22. We abstracted data on age, vaccination and prior infection history, disease severity assessed by oxygen requirement, hospital length of stay, and mortality via retrospective chart review.

Using this case definition based on the CDC criteria for severe disease, 67.5% of SARS-CoV-2 PCR positive hospitalized patients would not have met criteria for a COVID-19 hospitalization. These patients had significantly lower median age (44 years vs. 57 years), median hospital length of stay (2 days vs. 3 days), and in-hospital mortality (3.5% vs. 14%) (see Table). While unadjusted analysis did not show significant association between exposure to vaccine or prior infection and incidental infection (OR=0.79 [0.53-1.17], p=0.24), exposure to vaccine or prior infection was associated with incidental infection upon adjustment for age using logistic regression (OR=0.58 [0.38-0.89], p=0.01).

The high frequency of incidental COVID-19 infection among hospitalized patients detected using the case definition based on lack of oxygen requirement exceeds the rates reported in previous studies that used more stringent case definition based on complete absence of COVID-19 symptoms (2) or were performed during periods of the pandemic prior to the Omicron variant surge (3). However, the high frequency of incidental COVID-19 is very similar to measurements based on case definition of severe COVID-19 (6) or correlates such as administration of steroid treatment (5) during the Omicron surge. Given that non-severe COVID-19 infections not requiring supplementary oxygen can generally be treated on an outpatient basis, we propose that incidental infections in hospitalized patients should not be considered as COVID-19 hospitalizations in public health statistics used to inform the public or make policy decisions. One caveat is that patients with non-severe COVID-19 are hospitalized at a higher rate than patients without COVID-19 (4), which may reflect non-respiratory complications of COVID-19 including thrombosis or multisystem inflammation or exacerbation of underlying chronic diseases, although these complications are often difficult to attribute directly to COVID-19 in individual patients. An updated case definition resulting in more accurate measurement of COVID-19 hospitalizations will facilitate more effective health policy and trust with the public.

## Data Availability

All data produced in the present study are available upon reasonable request to the authors

## Patient Consent Statement

Patient consent is not applicable to this work, as patient data was collected via retrospective review of electronic medical records with the approval of the Institutional Review Board of the University of Southern California under Protocol HS-20-00880.

## Acknowledgments

We would like to thank Noah Wald-Dickler, MD, and Paul D. Holtom, MD, at LAC+USC Medical Center within the Department of Health Services of Los Angeles County, for their contributions to data collection.

---

**Table 1.** Characteristics of hospitalized patients with non-severe versus severe COVID-19 infection during the Omicron variant surge

|  | Non-Severe COVID-19 | Severe COVID-19 | p<sup>^</sup> |
| --- | --- | --- | --- |
| N | 312 | 150 |  |
| Age (median[IQR]) | 44 [30-59] | 57 [44-72] | <0.001 |
| Immunized* (%) | 186 (60.4) | 82 (54.7) | 0.24 |
| LOS<sup>#</sup> (median[IQR]) | 2 [1-4] | 3 [1-5] | <0.005 |
| Death | 11 (3.5) | 21 (14.0) | <0.001 |

\* “Immunized” is defined as having any exposure to SARS-CoV-2 vaccination or prior infection confirmed by PCR or antigen testing; 6 patients were missing data for either vaccination or prior infection
<sup>#</sup> Hospital length of stay among patients who survived to discharge
<sup>^</sup> p-value for Wilcoxon Rank Sum test (for age and LOS) or Pearson’s Chi-Squared test (for immunized and death)

